# Heterozygous *MMACHC* burden variants are associated with higher circulating vitamin B12 in the *All of Us* Research Program

**DOI:** 10.64898/2026.06.03.26354855

**Authors:** Ling Cai, Ralph J. DeBerardinis

## Abstract

Heterozygous carriers of autosomal recessive disease variants are conventionally considered unaffected, yet population-scale genomic datasets reveal subclinical carrier phenotypes. *MMACHC* encodes a cobalamin-processing protein whose biallelic loss causes cobalamin C deficiency, an inborn error of intracellular cobalamin metabolism. We performed an unbiased quantitative phenome-wide association screen in *All of Us* Research Program v8 to identify phenotypes associated with rare heterozygous *MMACHC* burden variants. Serum/plasma vitamin B12 was the top quantitative association. Carriers had higher circulating B12 than non-carriers in adjusted analyses, but also higher homocysteine, suggesting that elevated circulating B12 does not reflect improved intracellular cobalamin function. Carriers were less likely to fall below conventional B12 insufficiency thresholds, indicating a potential diagnostic blind spot. A pathway-wide rare-variant gene-burden (All-by-All) gene-burden analysis placed this finding in broader biological context. Burdens in genes related to circulating B12 binding or intestinal absorption were associated with lower circulating B12. In contrast, burdens in several genes involved in cellular delivery and intracellular cobalamin handling were associated with higher circulating B12. This step-specific directionality supports a model in which elevated circulating B12 can reflect impaired cellular handling and consequent systemic accumulation rather than improved cellular cobalamin availability. Because EHR-derived B12 is shaped by heterogeneous clinical and medication contexts, prospective carrier-enriched studies with standardized methylmalonic acid, homocysteine, diet, supplement, medication, comorbidity, and symptom ascertainment are needed to evaluate functional-marker-based screening.

## Main Text

Heterozygous carriers of autosomal recessive disease variants are often considered clinically unaffected, yet population biobanks can reveal measurable carrier phenotypes^1^. *MMACHC* encodes an intracellular cobalamin-processing protein. Biallelic *MMACHC* loss causes cobalamin C (cblC) disease, with methylmalonic acid (MMA) and homocysteine (Hcy) accumulation. Because *Mmachc* heterozygous mice show elevated MMA and Hcy^2^, we asked whether *MMACHC* heterozygosity leaves a detectable biochemical signature in humans.

We analyzed the *All of Us* Research Program v8 short-read whole-genome sequencing and linked electronic health record data^3^. *MMACHC* burden variants were defined as rare variants with ClinVar pathogenic/likely pathogenic (P/LP) annotation, predicted loss-of-function or splice-disrupting consequence, or missense consequence with REVEL score >0.6. Among 414,822 unrelated participants passing the analytic filters, 2,486 carried one *MMACHC* burden variant. We first performed a quantitative phenome-wide scan, adjusted for age, sex at birth, and genetic principal components across 174 laboratory concepts. Serum or plasma cobalamin/vitamin B12 was the top laboratory association, and the direction was unexpected: carriers had higher, not lower, circulating B12 (**Figure 1A-B**).

**Figure 1.**
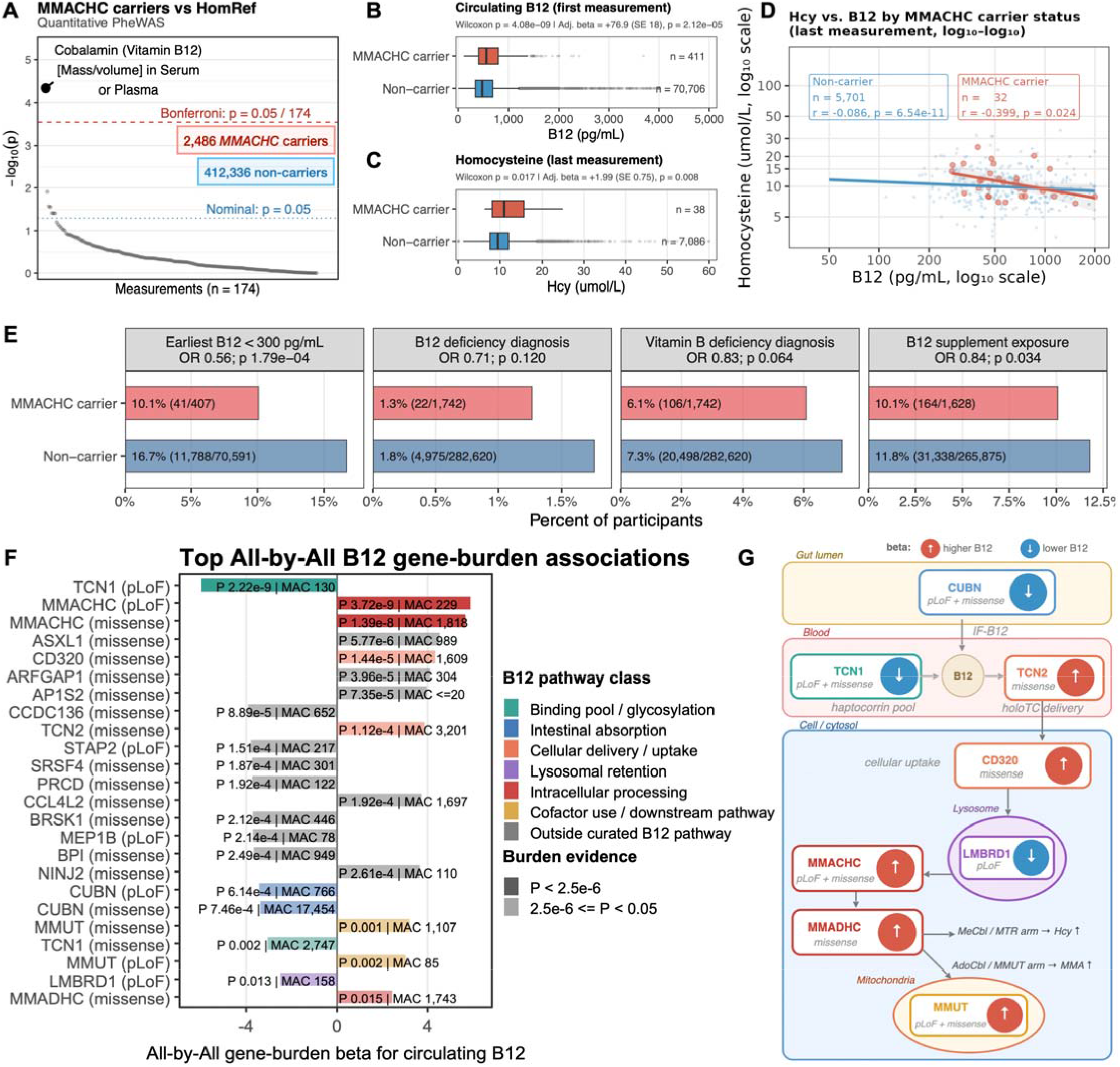
Elevated circulating B12 in *MMACHC* burden-variant carriers masks functional cobalamin insufficiency. (**A**) Quantitative PheWAS in the *All of Us* participants identifies serum/plasma cobalamin as the sole Bonferroni-significant association with *MMACHC* burden-variant carrier status. (**B-C**) Carriers have higher circulating B12 and Hcy than non-carriers, despite the smaller subset with available Hcy measurements. For repeated measurements, primary analyses used first B12 values to limit supplementation bias and last Hcy values to capture later functional status; alternative summaries gave similar associations. (**D**) An inverse relationship between circulating B12 and homocysteine is present in both carriers and non-carriers, based on last available measurements. (**E**) EHR-derived binary outcomes show carriers are less likely to be flagged below the 300 pg/mL insufficiency threshold and less likely to have a recorded B12 supplement exposure. ICD-coded deficiency diagnoses trend in the same direction. (**F**-**G**) Rare-variant gene-burden associations with circulating B12 from the *All of Us* All-by-All Browser vary systematically by cobalamin pathway position: burden in binding and absorption genes is associated with lower B12, while burden in genes mediating cellular delivery and intracellular processing is associated with higher B12, mirroring the *MMACHC* carrier direction.

Hcy was also higher among carriers in the subset with available testing (**Figure 1C**) and negatively associated with B12 levels in both carriers and non-carriers (**Figure 1D**), while MMA was too sparsely measured, and often below the reporting threshold, for a carrier-status analysis. Although B12 measurements were absent from UK Biobank, precluding direct validation, our finding aligns with a previous Icelandic and Danish GWAS for serum B12, which identified a rare P/LP *MMACHC* missense variant, R206Q, as one of the strongest genetic associations with increased serum B12 levels^4^.

These data suggest that higher circulating B12 may mask functional B12 deficiency in *MMACHC* burden-variant carriers (**Figure 1E**). Using the earliest available B12 measurement, which is less likely than later values to reflect subsequent supplementation or clinical intervention, *MMACHC* burden-variant carriers were significantly less likely than non-carriers to fall below the 300 pg/mL insufficiency threshold (10.1% vs. 16.7%; OR = 0.56; p = 1.79 × 10^−4^). A stricter 200 pg/mL deficiency threshold showed the same direction (OR = 0.57) but did not reach significance, likely reflecting limited power given the small number of deficient carriers. ICD-coded B12 deficiency diagnoses trended in the same direction but did not reach significance, likely reflecting the lower sensitivity of diagnosis codes compared to direct laboratory thresholds. Carriers were also modestly less likely to have a recorded B12 supplement exposure (OR = 0.84; p = 0.034), suggesting that the lower rate of threshold-based flagging propagates into reduced clinical recognition and treatment. Hence, circulating B12 alone may be an unreliable reassurance marker in *MMACHC* carriers, and functional markers such as Hcy and MMA may better capture carrier risk.

To place the *MMACHC* result in pathway context, we examined aggregate All-by-All Browser B12 gene-burden results across cobalamin-related genes (**Figures 1F-G**). The sign of association varied by pathway position. Burden variation in binding or absorption genes, including *TCN1* and *CUBN*, was associated with lower circulating B12, whereas burden variation in post-uptake and intracellular cobalamin-handling genes, including *TCN2, CD320, MMACHC, MMADHC*, and *MMUT*, tended to be associated with higher circulating B12. This step-specific directionality argues against a nonspecific artifact and instead supports a model in which elevated circulating B12 can reflect impaired cellular handling and consequent systemic accumulation, rather than improved intracellular cobalamin availability.

The analysis is observational, EHR laboratory testing was not standardized, and Hcy measurements were available only in a clinically selected subset. Nevertheless, the results identify a reproducible carrier-associated B12 phenotype with a plausible diagnostic blind spot — one that may be compounded in carriers who also carry additional B12-relevant stressors such as South Asian ancestry, nicotine use, digestive disorders, pregnancy, or low dietary B12 intake^5^. Recruiting relatives of cblC probands identified through newborn screening or metabolic clinics would enrich for familial *MMACHC* pathogenic alleles and enable prospective paired measurement of B12, Hcy, and MMA with standardized dietary and medication ascertainment, providing the targeted follow-up needed to determine whether heterozygous *MMACHC* burden variants modify functional B12 requirements, pregnancy outcomes, or neurological risk.

### Declaration of generative AI and AI-assisted technologies in the manuscript preparation process

During the preparation of this work, the authors used ChatGPT, Biomni, and Codex to support manuscript organization, language refinement, conceptual figure planning, and analysis-code drafting or debugging. After using these tools, the authors reviewed and edited the content and code as needed, independently verified the scientific content, references, analyses, and figures, and take full responsibility for the content of the publication.

## Data Availability

This study used the All of Us Research Program Controlled Tier Dataset v8. All participant-level data were accessed and analyzed within the All of Us Researcher Workbench and cannot be redistributed by the authors. The All of Us data are available to authorized researchers through the Researcher Workbench after completion of the required access process and data-use agreements. Exported materials from this study were limited to aggregate summaries, masked tables, analysis scripts, and figures consistent with All of Us data dissemination requirements.

## Notes

### Competing Interest Statement

RJD is a founder and advisor at Atavistik Bioscience and an advisor at Vida Ventures, Agios Pharmaceuticals, Illumina, and Faeth Therapeutics.

### Author Declarations

The Institutional Review Board of the All of Us Research Program gave ethical approval for this work.

### Summary of Updates

Downsized from Research Article to Research Letter format.

